# Development and external validation of a nomogram for predicting re-fracture after percutaneous kyphoplasty: A two-center study

**DOI:** 10.64898/2026.11.19.26368352

**Authors:** Weixiang Wu, Wenxuan Cui

## Abstract

**Background:** To identify independent risk factors for subsequent vertebral re-fracture after percutaneous kyphoplasty (PKP) in patients with osteoporotic vertebral compression fracture (OVCF) and to develop and externally validate a predictive nomogram for individualized risk assessment.

**Methods:** This two-center retrospective study enrolled 1,720 OVCF patients who underwent PKP between January 2023 and April 2025. The training cohort (n=963) and internal validation cohort (n=413) were derived from Beijing LUHE Hospital, while the external validation cohort (n=344) was from the other participating center. The primary outcome was postoperative re-fracture confirmed by imaging within a minimum 12-month follow-up. To find independent risk variables in the training cohort, multivariate logistic regression analysis was performed after potential predictors were screened using least absolute shrinkage and selection operator (LASSO) regression. Based on these variables, a nomogram was created. Receiver operating characteristic (ROC) curves, calibration curves, and decision curve analysis (DCA) were used to evaluate model performance for each of the three cohorts.

**Results:** Five independent factors of re-fracture were found using multivariate logistic regression: advanced age (≥65 years) (OR=2.58, 95% CI: 1.81-3.67), body mass index (BMI) ≥18.5 (protective, OR=0.32, 95% CI: 0.21-0.49), bone mineral density (BMD) (OR=0.13, 95% CI: 0.08-0.20), osteocalcin (BGP) (OR=1.16, 95% CI: 1.09-1.22), and Cobb angle correction (OR=1.36, 95% CI: 1.21-1.53). The nomogram demonstrated good discrimination in the training cohort (AUC=0.810, 95% CI: 0.779-0.841), internal validation cohort (AUC=0.801, 95% CI: 0.732-0.870), and external validation cohort (AUC=0.809, 95% CI: 0.750-0.867). DCA verified net benefit over a threshold probability range of 0-0.6, and calibration curves demonstrated good agreement between projected and observed probabilities.

**Conclusion:** The externally validated nomogram, incorporating age, BMI, BMD, BGP, and Cobb angle correction, provides a robust and practical tool for individualized prediction of re-fracture risk after PKP in OVCF patients, which could facilitate risk-stratified postoperative management and potentially improve patient outcomes.

## Introduction

Osteoporotic vertebral compression fracture (OVCF) are becoming more common as the world’s population ages ^1^. Patients usually have severe back pain, kyphotic deformity, short height and limited mobility, which can considerably lower patients’ quality of life and place a considerable financial strain on families and society ^2^. Although OVCF have traditionally been regarded as a senile disease, more and more young patients have been diagnosed with this disease in clinical practice. It is worth noting that the etiology, bone condition and complication characteristics of these young patients are often significantly different from those of elderly patients ^3^.

The treatment of OVCF mainly includes two methods: Both surgical and conservative methods of treatment. Although conservative treatment is still the main choice, it has significant disadvantages, including slow analgesic effect, long treatment cycle and long bed rest. This puts patients prone to problems such as pressure sores, respiratory and urinary tract infections, and constipation ^4,5^. For OVCF patients with poor response to conservative treatment, percutaneous kyphoplasty (PKP) has become the mainstream surgical intervention method ^6^. PKP restores the height of the vertebral body through balloon expansion and injects bone cement to provide mechanical stability. The operation can quickly relieve pain, rebuild the stability of the spine and promote early bed activity, thus effectively reducing the risk of long-term bed rest ^7^. However, despite the satisfactory therapeutic effect shown in short-term follow-up, the long-term complications of PKP, especially postoperative fractures, are attracting more and more clinical attention ^8^. A large number of studies have shown that during different follow-up periods, the incidence of postoperative fractures after PKP in patients with OVCF can exceed 15% ^9-11^. Such re-fracture events may not only cause a vicious circle of pain recurrence and function decline, but also aggravate spondylosis, posing a serious threat to the long-term prognosis of patients ^12,13^.

At present, refractures after PKP have been widely regarded as a complex clinical problem driven by a variety of factors. Age, sex, bone mineral density (BMD), bone cement leakage, the number of treated vertebrae, and a history of diabetes are just a few of the potential risk variables that have been found in earlier study, but the findings are still conflicting ^14-17^. In addition, most studies only focus on isolated single factors and lack a practical clinical tool that can systematically integrate multiple independent predictors and provide accurate risk quantification for individual patients. Nomogram is a visual prediction tool based on regression models, which transforms complex mathematical formulas into intuitive graphical interfaces. By integrating multiple predictor variables, it enables clinicians to estimate the probability of a specific clinical event ^18^. In recent years, this tool has shown remarkable practical value in oncology and other fields, and is gradually being applied to the construction of orthopedic prognosis prediction models. However, there is still a lack of reliable nomogram models to predict the risk of re-fracture in patients with OVCF after PKP surgery.

Therefore, this study aims to systematically identify the independent risk factors for secondary fractures after PKP in patients with OVCF, and build a reliable predictive nomogram based on these factors. The nomogram proposed by our two medical institutions is expected to become a practical clinical tool that can accurately stratify risks and help make personalized postoperative management decisions for different patient groups.

## Methods

### Patients and study design

This two-center, retrospective study enrolled 1,720 patients with OVCF who underwent PKP between January 2023 and April 2025. The training cohort (n = 963) and internal validation cohort (n = 413) were derived from Beijing LUHE Hospital, while the external validation cohort (n = 344) was derived from the Beijing Tongzhou District Hospital of Integrated Traditional Chinese and Western Medicine. Inclusion criteria: (1) Confirmed as a new osteoporotic vertebral compression fracture (OVCF) according to clinical manifestations and imaging examination ^19^; Received PKP treatment for the first time; (3) No history of lumbar surgery; (4) Postoperative follow-up time of more than 12 months; (5) The bed information is complete. Exclusion criteria: (1) pathological fractures caused by malignant tumors, infections, etc.; (2) complications that seriously affect bone metabolism, such as hyperparathyroidism or rheumatoid arthritis; (3) neurological diseases that may lead to abnormal gait or significantly increase the risk of falling, such as Parkinson’s disease.

This study adhered to the principles of the Declaration of Helsinki, as revised in 2013. The research protocol received ethical approval from the Ethics Committee of Beijing LUHE Hospital (reference number 2025LH-ICY-033) and from the ethics boards of all collaborating institutions. Given the retrospective nature of the data analysis, informed consent was waived by all approving committees.

### Follow-up

All patients receive a follow-up evaluation every three months after the operation, which is carried out through outpatient re-examination or telephone inquiry. The follow-up period of all patients was fixed at 12 months after PKP, and it was evaluated whether there were re-fractures within those 12 months. The main outcome indicator is postoperative refracture, which is defined as imaging examination (X-ray or computed tomography) showing new evidence of vertebral compression fracture, accompanied by corresponding clinical symptoms. The follow-up cutoff date was April 2026, by which all included patients had completed the fixed 12-month follow-up.

### Data collection

Potential risk factors for re-fracture were extracted from hospital records. Demographic characteristics included age, sex, body mass index (BMI), alcohol consumption (>15 g/day), and smoking status (yes/no). Comorbidities recorded included hypertension, diabetes, and hyperlipidemia. Injury mechanism was classified as high-energy (e.g., fall from >4 feet, traffic accident, direct blow) or low-energy (fall from standing or sitting position) ^20^. Before surgery, dual-energy X-ray absorptiometry was used to evaluate bone mineral density (BMD). Surgical parameters collected included number of treated vertebrae, and bone cement leakage (yes/no) on postoperative radiographs. Radiographic measurements included Cobb angle correction (postoperative change in degrees). The Spinal Deformity Index (SDI) was calculated as the sum of deformity scores (0-3 per vertebra) for pre-existing fractures from T4 to L4 ^21^. Inter-observer reliability for SDI was assessed in a random sample of 100 patients by two spine surgeons (intraclass correlation coefficient = 0.89; 95% CI: 0.84-0.93). Before surgery, venous blood samples were taken from each patient while they were fasting. All laboratory analyses were performed uniformly by the hospital’s central clinical laboratory. The following indicators were measured: osteocalcin (BGP), calcium, phosphorus.

### Statistical analysis

All statistical analysis is carried out using SPSS 26.0 and R 4.5.1 software. Frequency and percentage are used to represent classification variables, whereas mean ± standard deviation or median (interquartile range) are used to express continuous data. For the baseline comparison between three cohorts (training cohort, internal validation cohort and external validation cohort), after evaluating the normality through the Shapiro-Wilk test and evaluating the variance uniformity through the Levene’s test, the chi-square test (or Fisher’s exact test) is used to compare the differences between groups of the classification variables, and the continuous variables one-way analysis of variance or Kruskal-Wallis H test is used for the quantity. In this two-center study, patients from one of the centers were randomly assigned to the training cohort (n=963) and the internal validation cohort (n=413) at a ratio of 7:3, while patients from the other center were assigned to the external test cohort (n=344). In order to reduce the dimension and avoid overfitting, we applied the least absolute shrinkage and selection operator (LASSO) regression to the candidate variables using the training cohort. The R language’s glmnet package is used for LASSO analysis, and 10-fold cross-validation using the least mean square error criterion yields the ideal parameter (λ). Variables having non-zero coefficients at the optimal λ value are reserved as potential predictors. Subsequently, multivariable logistic regression analysis was carried out on the variables selected by LASSO in the training cohort to determine the independent risk factors for re-fracture after PKP in OVCF patients. The odds ratio (OR) and its 95% confidence interval (CI) were calculated. Based on the independent risk factors determined by the multivariable logistic regression analysis, a nomogram model for predicting the risk of individual refracture was constructed using the rms package in R 4.5.1. The model performance was evaluated in the training cohort, and internal validation was carried out in the internal validation cohort, and external validation was carried out in the external validation cohort. Evaluate the distinguishing ability of the model by drawing the receiver operating characteristic (ROC) curve and calculating the area under the curve (AUC) of the three cohorts. Draw the calibration curve to evaluate the calibration of the model in each cohort. Decision curve analysis (DCA) was carried out to evaluate the net benefit of the model under different threshold probabilities in the training cohort, internal validation cohort and external validation cohort. In this study, P < 0.05 is considered to be statistically significant.

## Results

### Characteristics of patients

This study had 1,720 patients in total: 963 in the training cohort, 413 in the internal validation cohort, and 344 in the external validation cohort. The numbers of re-fracture in the training, internal validation, and external validation cohorts were 214 (22.2%), 83 (20.1%), and 72 (20.9%), respectively. For the distributions of age, sex, BMI, alcohol consumption, smoking status, injury mechanism, bone cement leakage, hypertension, diabetes, hyperlipidemia, number of treated vertebrae, BMD, BGP, calcium, phosphorus, cobb angle correction, and SDI were comparable among the three cohorts, with all *P* > 0.05. The characteristics of the patients across the three cohorts are summarized in **Table 1**.

**Table 1.**
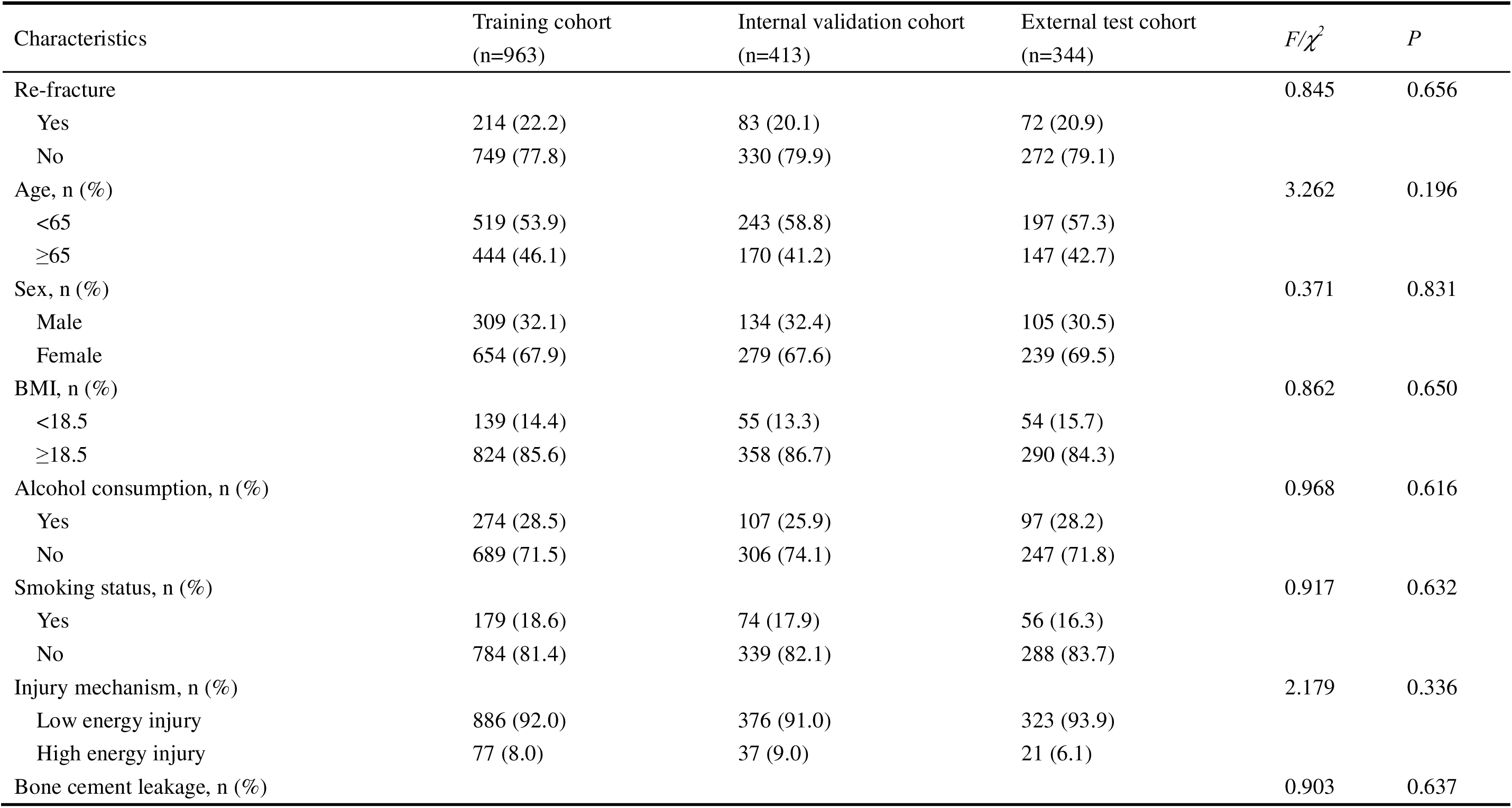
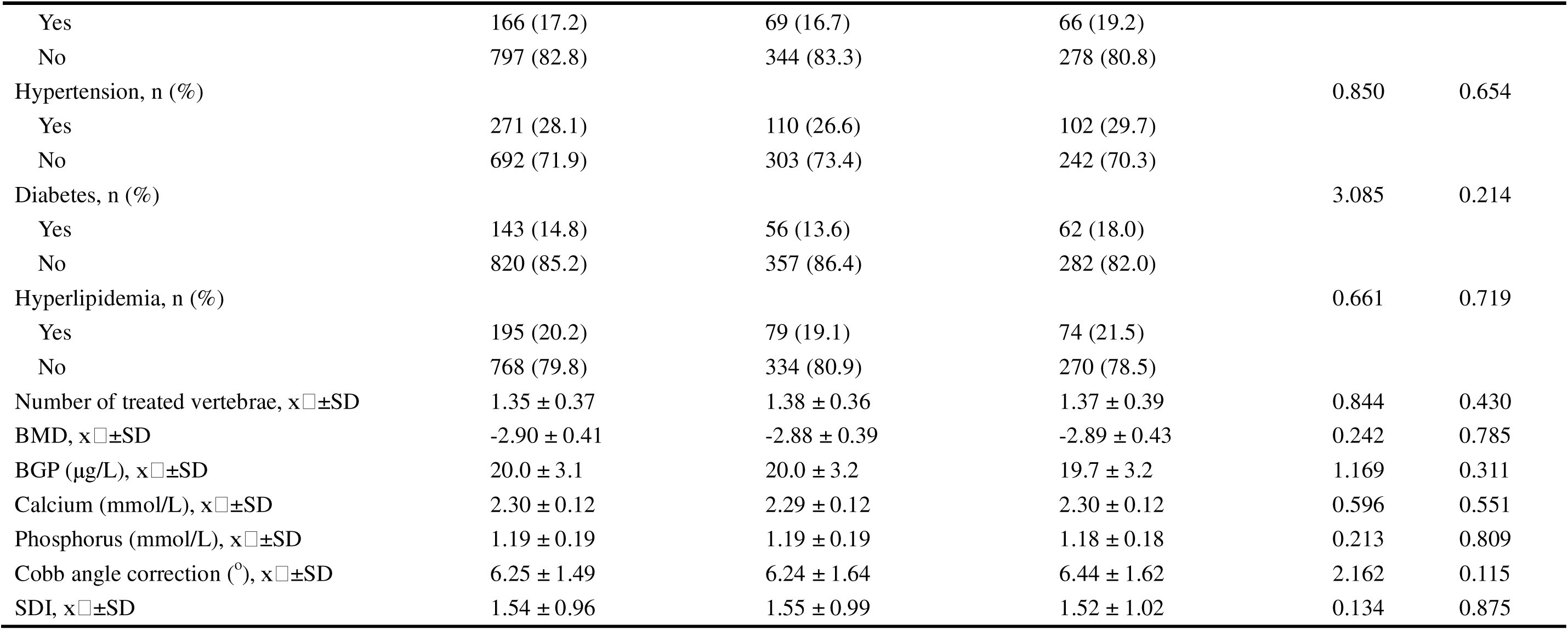
Characteristics of patients (n=1720).

| Characteristics | Training cohort<br>(n=963) | Internal validation cohort<br>(n=413) | External test cohort<br>(n=344) | *F*/*χ*<sup>2</sup> | *P* |
|---|---|---|---|---|---|
| Re-fracture |  |  |  | 0.845 | 0.656 |
| Yes | 214 (22.2) | 83 (20.1) | 72 (20.9) |  |  |
| No | 749 (77.8) | 330 (79.9) | 272 (79.1) |  |  |
| Age, n (%) |  |  |  | 3.262 | 0.196 |
| <65 | 519 (53.9) | 243 (58.8) | 197 (57.3) |  |  |
| ≥65 | 444 (46.1) | 170 (41.2) | 147 (42.7) |  |  |
| Sex, n (%) |  |  |  | 0.371 | 0.831 |
| Male | 309 (32.1) | 134 (32.4) | 105 (30.5) |  |  |
| Female | 654 (67.9) | 279 (67.6) | 239 (69.5) |  |  |
| BMI, n (%) |  |  |  | 0.862 | 0.650 |
| <18.5 | 139 (14.4) | 55 (13.3) | 54 (15.7) |  |  |
| ≥18.5 | 824 (85.6) | 358 (86.7) | 290 (84.3) |  |  |
| Alcohol consumption, n (%) |  |  |  | 0.968 | 0.616 |
| Yes | 274 (28.5) | 107 (25.9) | 97 (28.2) |  |  |
| No | 689 (71.5) | 306 (74.1) | 247 (71.8) |  |  |
| Smoking status, n (%) |  |  |  | 0.917 | 0.632 |
| Yes | 179 (18.6) | 74 (17.9) | 56 (16.3) |  |  |
| No | 784 (81.4) | 339 (82.1) | 288 (83.7) |  |  |
| Injury mechanism, n (%) |  |  |  | 2.179 | 0.336 |
| Low energy injury | 886 (92.0) | 376 (91.0) | 323 (93.9) |  |  |
| High energy injury | 77 (8.0) | 37 (9.0) | 21 (6.1) |  |  |
| Bone cement leakage, n (%) |  |  |  | 0.903 | 0.637 |
| Yes | 166 (17.2) | 69 (16.7) | 66 (19.2) |  |  |
| No | 797 (82.8) | 344 (83.3) | 278 (80.8) |  |  |
| Hypertension, n (%) |  |  |  | 0.850 | 0.654 |
| Yes | 271 (28.1) | 110 (26.6) | 102 (29.7) |  |  |
| No | 692 (71.9) | 303 (73.4) | 242 (70.3) |  |  |
| Diabetes, n (%) |  |  |  | 3.085 | 0.214 |
| Yes | 143 (14.8) | 56 (13.6) | 62 (18.0) |  |  |
| No | 820 (85.2) | 357 (86.4) | 282 (82.0) |  |  |
| Hyperlipidemia, n (%) |  |  |  | 0.661 | 0.719 |
| Yes | 195 (20.2) | 79 (19.1) | 74 (21.5) |  |  |
| No | 768 (79.8) | 334 (80.9) | 270 (78.5) |  |  |
| Number of treated vertebrae, x□±SD | 1.35 ± 0.37 | 1.38 ± 0.36 | 1.37 ± 0.39 | 0.844 | 0.430 |
| BMD, x□±SD | -2.90 ± 0.41 | -2.88 ± 0.39 | -2.89 ± 0.43 | 0.242 | 0.785 |
| BGP (μg/L), x□±SD | 20.0 ± 3.1 | 20.0 ± 3.2 | 19.7 ± 3.2 | 1.169 | 0.311 |
| Calcium (mmol/L), x□±SD | 2.30 ± 0.12 | 2.29 ± 0.12 | 2.30 ± 0.12 | 0.596 | 0.551 |
| Phosphorus (mmol/L), x□±SD | 1.19 ± 0.19 | 1.19 ± 0.19 | 1.18 ± 0.18 | 0.213 | 0.809 |
| Cobb angle correction (<sup>o</sup>), x□±SD | 6.25 ± 1.49 | 6.24 ± 1.64 | 6.44 ± 1.62 | 2.162 | 0.115 |
| SDI, x□±SD | 1.54 ± 0.96 | 1.55 ± 0.99 | 1.52 ± 1.02 | 0.134 | 0.875 |

### Multivariate analysis and independent risk factors

LASSO regression was used to evaluate the data. As the penalty parameter λ grows, **Figure 1** shows how the coefficients of all independent variables fall toward zero until all coefficients are reduced to zero. The optimal λ=0.034 was determined by 10-fold cross-validation, and the binomial deviance reached its minimum when five variables (age, BMI, BMD, BGP, Cobb angle correction) were included in the model. A multivariable logistic regression model was constructed with the occurrence of postoperative re-fracture as the dependent variable and the factors identified through LASSO regression as independent variables, to further identify significant predictive variables. Details of variable coding are provided in **Supplementary Table 1**.

**Figure 1.**
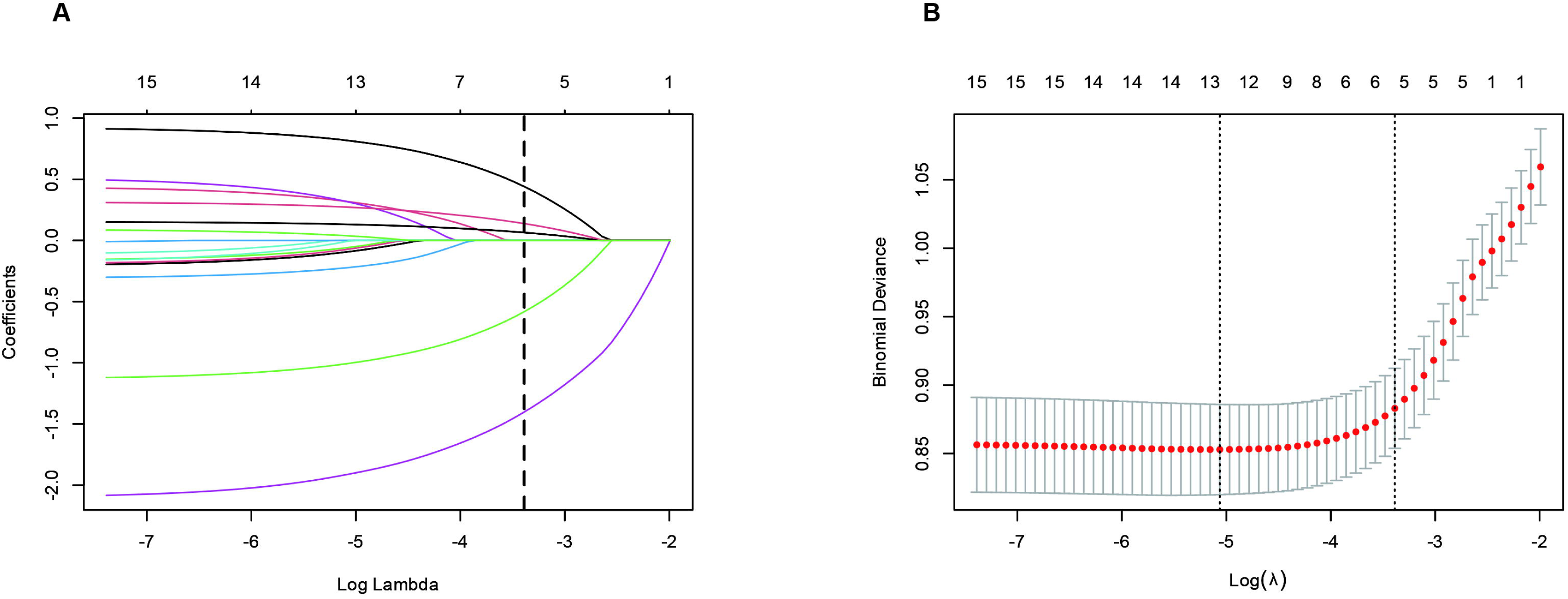
Characteristics of coefficient variation in least absolute shrinkage and selection operator (LASSO) regression and analysis of binomial deviance plots.

In order to determine the independent risk factors of re-fracture after PKP, a multivariable logistic regression analysis was carried out on the training cohort (n=963). The results are shown in Table 2. The analysis results show that there are several factors that are independently related to the occurrence of another fracture. Advanced age (≥65 years) was identified as a significant risk factor, with an OR of 2.58 (95% CI: 1.81-3.67, *P* < 0.05) compared to the younger age group (<65 years). A higher BMI (≥18.5) emerged as a protective factor, significantly reducing the risk of re-fracture (OR = 0.32, 95% CI: 0.21-0.49, *P* < 0.05). Additionally, lower BMD was strongly associated with an increased risk of re-fracture (OR = 0.13, 95% CI: 0.08-0.20, *P* < 0.05). Elevated levels of BGP were also found to be an independent risk factor (OR = 1.16, 95% CI: 1.09-1.22, *P* < 0.05). Finally, a greater degree of Cobb angle correction was identified as a significant predictor of re-fracture (OR = 1.36, 95% CI: 1.21-1.53, *P* < 0.05).

**Table 2.** Results of multivariate logistic regression in training cohort (n=963).

| **Characteristics** | ***β*** | **SE** | **OR (95% CI)** | ***P*** |
|---|---|---|---|---|
| Age |  |  |  |  |
| <65 |  |  | 1 (References) | - |
| ≥65 | 0.946 | 0.180 | 2.58 (1.81-3.67) | <0.001 |
| BMI |  |  |  |  |
| <18.5 |  |  | 1 (References) | - |
| ≥18.5 | -1.138 | 0.219 | 0.32 (0.21-0.49) | <0.001 |
| BMD | -2.068 | 0.235 | 0.13 (0.08-0.20) | <0.001 |
| BGP | 0.146 | 0.029 | 1.16 (1.09-1.22) | <0.001 |
| Cobb angle correction | 0.306 | 0.060 | 1.36 (1.21-1.53) | <0.001 |

### Construction and validation of the re-fracture nomogram models

We created a nomogram model to predict patients’ postoperative re-fracture based on the independent factors found by the multivariable logistic regression analysis (**Figure 2**). The area under the ROC curve was 0.810 (95% CI: 0.779-0.841) for the training cohort, 0.801 (95% CI: 0.732-0.870) for the internal validation cohort, and 0.809 (95% CI: 0.750-0.867) for the external validation cohort (**Figure 3**). The calibration curves showed that the actual observed outcomes and the anticipated probabilities agreed well (**Figure 4**). The prediction model for patients offered a net benefit within a threshold probability range of 0-0.6, according to decision curve analysis (**Figure 5**).

**Figure 2.**
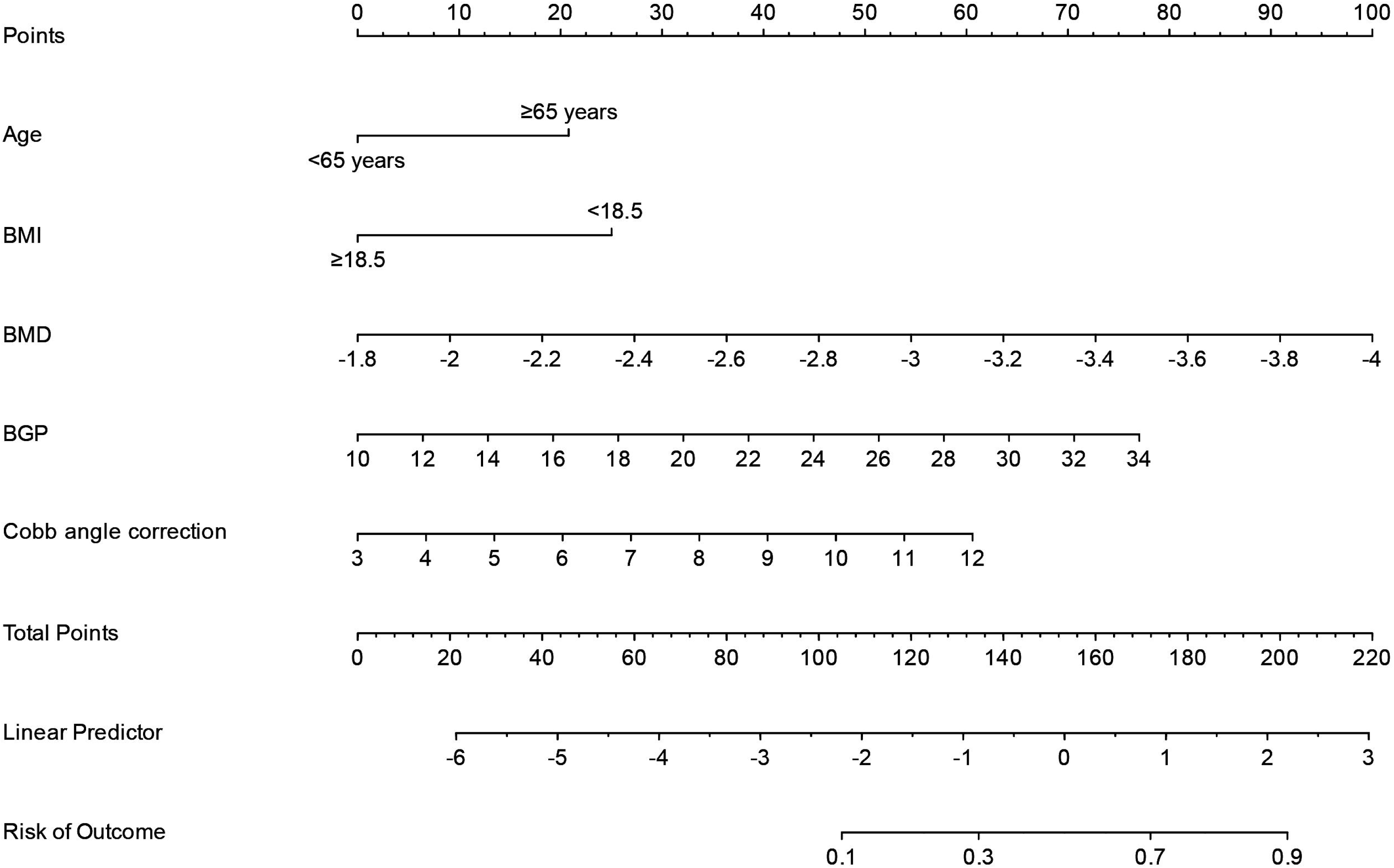
Nomogram for predicting re-fracture.

**Figure 3.**
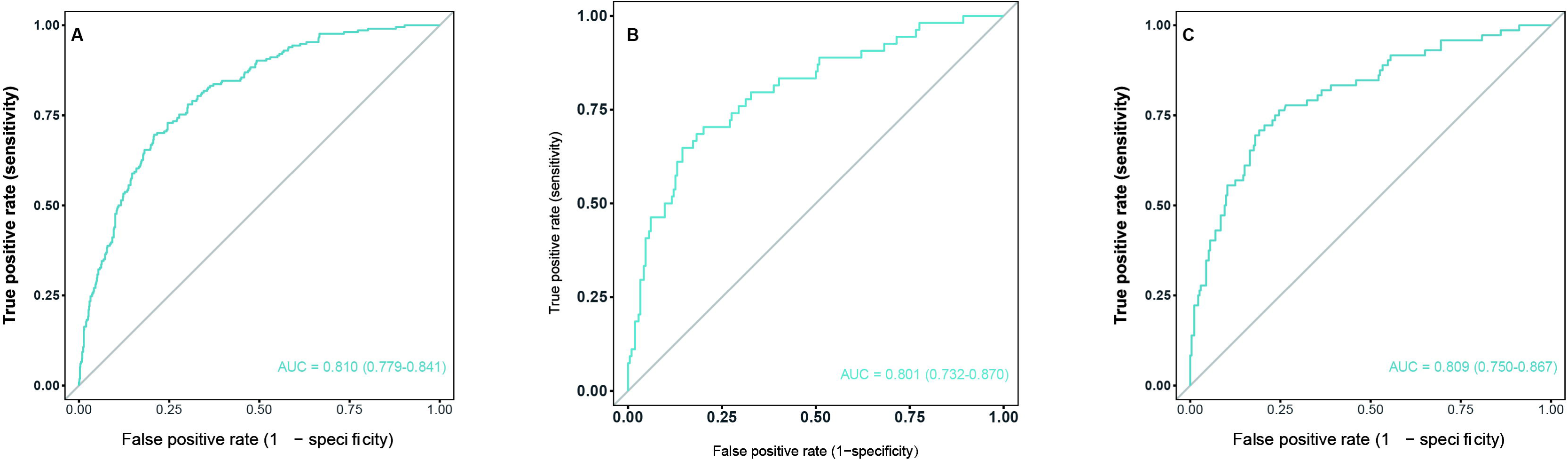
ROC curves for the nomogram. A, training cohort; B, internal validation cohort; C, external test cohort.

**Figure 4.**
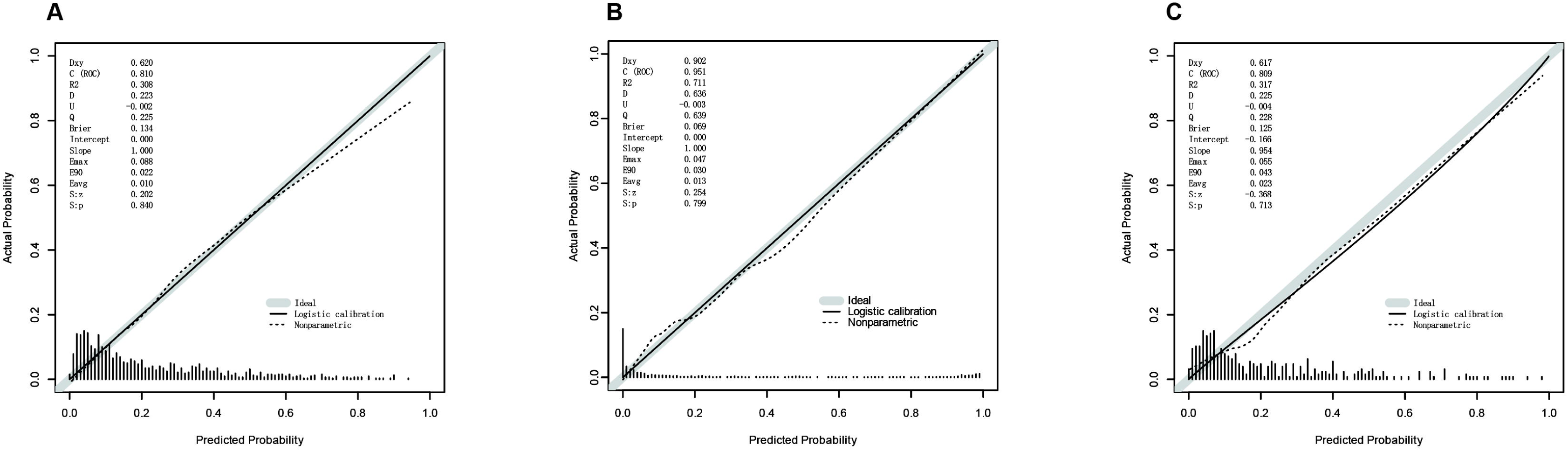
Calibration curve for the nomogram. A, training cohort; B, internal validation cohort; C, external test cohort. The vertical coordinate was the actual probability of re-fracture. The Ideal line was the diagonal dashed line, representing the predicted probability equal to the actual probability under the optimal condition. The Apparent line and the Bias-corrected line, respectively, represent the predicted probability and the actual probability of the nomogram prediction model constructed in this study under the present real conditions. The closer these two lines were to the Ideal line (diagonal dashed line), the better the nomogram prediction model was, and the better its calibration was.

**Figure 5.**
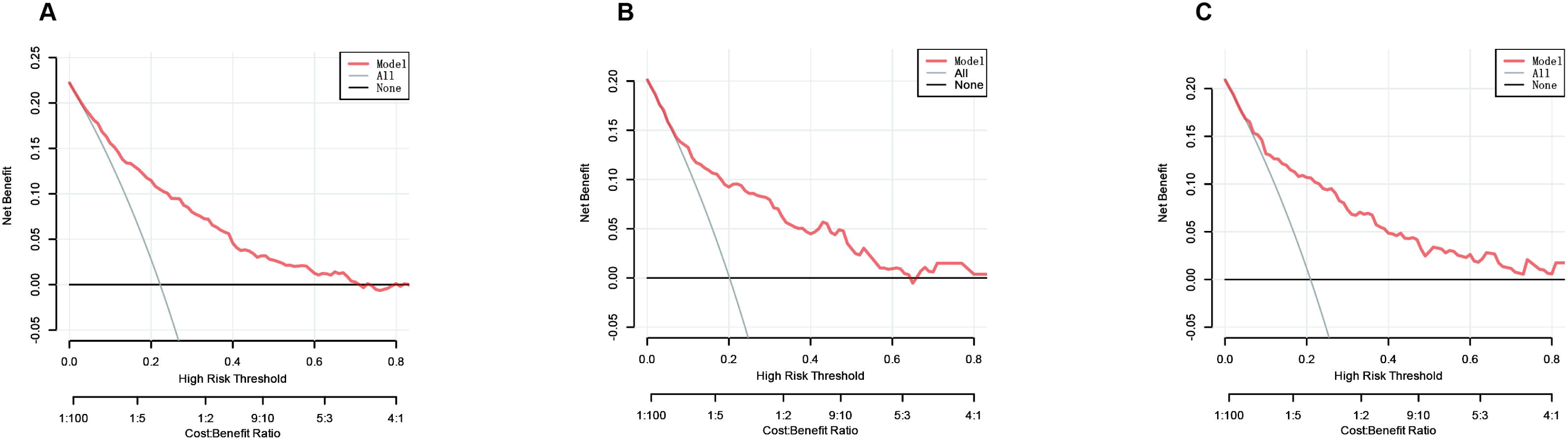
Decision curve for the nomogram. A, training cohort; B, internal validation cohort; C, external test cohort. The red line (Nomogram model) represented the overall benefit if screening was performed based on the nomogram prediction model constructed for this study. The gray line (ALL) represented the overall benefit of considering all participants are assumed to experience a re-fracture. The gray-black line (None) represented the assumption that none of the participants experience a re-fracture, and thus, no action was taken. No cost was incurred, but there was no clinical benefit either.

## Discussion

This two-center retrospective cohort study successfully developed and externally validated a nomogram for predicting re-fracture after PKP in patients with OVCF. Based on the training cohort, the multivariate logistic regression analysis identified five independent predictors: advanced age (≥65 years), lower BMI (<18.5), lower BMD, BGP levels, and greater Cobb angle correction. By comprehensively considering these factors, our nomogram shows good discrimination ability in both internal and external validation cohorts, with AUC values of 0.801 and 0.809 respectively. These research results provide a practical and reliable clinical tool for individualized risk stratification and postoperative management.

This study not only confirms several established risk factors again, but also provides new insights. Advanced age (≥65 years old) has been confirmed as a significant independent risk factor (OR = 2.58), which is consistent with the widely accepted view that aging is related to bone microstructure degradation, bone metabolism slowdown and increased risk of fall ^15,22,23^. This finding emphasizes that age remains a basic indicator of cumulative bone vulnerability, and its strong predictive value in this model confirms the rationality of continuing to include this indicator in the risk assessment. The protective effect of higher BMI (OR = 0.32) and higher bone density (BMD, OR = 0.13) further confirms the key role of systemic bone health and mechanical support in the prevention of subsequent fractures. Lower BMI (which usually suggests sarcopenia) weakens the patient’s overall mechanical toughness and soft tissue buffering ability ^24^, while lower BMD directly reflects a decrease in bone mineral content and structural integrity ^25^. These results strongly support the need to implement comprehensive anti-osteoporotic and nutritional optimization, including weight-bearing exercise and protein supplementation, to reduce the risk of re-fractures.

A novel and remarkable finding is that elevated preoperative BGP levels are identified as an independent risk factor (OR = 1.16). As a marker of bone formation, BGP is traditionally considered to reflect the activity of osteoblasts. However, the phenomenon of elevated levels in the case of re-fractures is quite intriguing ^26^. A reasonable explanation is that the increase in BGP levels in these patients may indicate that they are in a state of high bone turnover, in which dissociated or dysregulated bone remodeling will lead to the formation of poor-quality bone with impaired mechanical properties ^27^. In addition, this elevation may also reflect a systemic stress response or secondary hyperparathyroidism caused by vitamin D deficiency, although we lack corresponding biochemical data to confirm this mechanism ^28^. This finding emphasizes that bone metabolic markers (not just static bone density) can provide valuable dynamic prognosis information. Another important factor leading to the risk of re-fracture is the greater amplitude of Cobb angle correction (OR = 1.36). This shows that although the degree of posterior correction achieved in surgery is conducive to spinal alignment, it may paradoxically increase the mechanical load of the adjacent vertebra or the therapeutic segment itself. This may be due to changes in the biomechanics and stress distribution of the spine, because excessive correction may concentrate the force on the bone cement-vertebral interface or adjacent vertebrae, making it more likely to have new compression fractures ^29-31^. This finding advocates a more cautious surgical strategy, emphasizes "optimal correction" rather than "maximum correction", and highlights the importance of postoperative biomechanical assessment.

In terms of predictive performance, our nomogram shows strong discrimination ability in all three cohorts: in the training cohort, AUC is 0.810 (95% CI: 0.779-0.841), and in the internal validation cohort, it is 0.801 (95% CI: 0.732-0.870), 0.809 (95% CI: 0.750-0.867) in the external validation cohort. The higher AUC value in the external validation cohort is particularly noteworthy, because it confirms the generalization ability of the model outside the derivation center ^32^. The calibration curve further shows that there is good consistency between the predicted probability and the observation results, and the decision curve analysis (DCA) confirms that the model has a positive net benefit within a wide threshold probability range (0-0.6). These results comprehensively show that our nomogram can not only accurately stratify patients by risk, but also have practical clinical practical value, enabling clinical clinical monitoring or targeted preventive interventions.

This study has several important advantages, including two-center design, large sample size and the inclusion of external validation cohort. However, it should also be admitted that there are some limitations. First of all, its retrospective nature may introduce selection bias, although external verification can help alleviate this problem. Secondly, although we have determined that elevated BGP is a predictor, the lack of postoperative continuous measurement data limits our ability to track the dynamic changes of bone metabolism markers and their relationship with fracture healing and long-term prognosis. Third, this study did not collect the use of preoperative anti-osteoporotic, exposure to glucocorticoids and vitamin D levels, nor did it evaluate the compliance of postoperative anti-osteoporotic. These factors may significantly affect bone quality and the risk of re-fractures. In the future, it is necessary to carry out prospective multi-center research, adopt a standardized postoperative management scheme and conduct longitudinal biomarker evaluation to further verify and optimize our prediction model.

## Conclusion

In summary, this two-center study found that advanced age (≥65 years old), low BMI (<18.5), low bone density, high BGP levels and greater Cobb angle correction amplitude are independent risks of fractures after PKP in patients with OVCF. Factors. Based on these factors, we have successfully developed and carried out external verification to establish a practical nomogram, which has good distinction, calibration and clinical practicality. This tool can facilitate individualized risk assessment and guide targeted postoperative prevention strategies, potentially improving long-term outcomes in this patient population.

## Declarations

### Authors’ contributions

Weixiang Wu: Funding acquisition, Writing - Original Draft, Writing - Review & Editing. Wenxuan Cui: Conceptualization, Methodology

## Acknowledgements

All authors thank Beijing LUHE Hospital and Beijing Tongzhou District Hospital of Integrated Traditional Chinese and Western Medicine for providing the clinical data.

## Data availability

The datasets used and analysed during the current study are not publicly available due to the authors do not have permission to share data, but are available from the corresponding author on reasonable request.

