## supplementary materials for "Development and external validation of a nomogram for predicting re-fracture after percutaneous kyphoplasty: A two-center study"

**Supplementary Table 1** Variable assignment.

| Variable | Assignment |
| --- | --- |
| Age | 0=<65; 1=≥65 |
| Gender | 0=Female; 1=Male |
| BMI | 0=<18.5; 1=≥18.5 |
| Alcohol consumption | 0=No; 1=Yes |
| Smoking status | 0=No; 1=Yes |
| Injury mechanism | 0=Low energy injury; 1=High energy injury |
| Bone cement leakage | 0=No; 1=Yes |
| Hypertension | 0=No; 1=Yes |
| Diabetes | 0=No; 1=Yes |
| Hyperlipidemia | 0=No; 1=Yes |
| Number of treated vertebrae | Original value substitution |
| BMD | Original value substitution |
| BGP (ug/L) | Original value substitution |
| Calcium (mmol/L) | Original value substitution |
| Phosphorus (mmol/L) | Original value substitution |
| Cobb angle correction (^o^) | Original value substitution |
| SDI | Original value substitution |
